# Different Covid-19 Outcomes Among Systemic Rheumatic Diseases: A Nation-wide Cohort Study

**DOI:** 10.1101/2022.03.11.22271887

**Authors:** Vasiliki-Kalliopi Bournia, George E. Fragoulis, Panagiota Mitrou, Konstantinos Mathioudakis, Anastasios Tsolakidis, George Konstantonis, Ioulia Tseti, Georgia Vourli, Maria G. Tektonidou, Dimitrios Paraskevis, Petros P. Sfikakis

## Abstract

**Background:** Nationwide data at a country level on Covid-19 in unvaccinated patients with rheumatoid arthritis (RA), ankylosing spondylitis (AS), psoriatic arthritis (PsA), systemic lupus erythematosus (SLE) and systemic sclerosis (SSc) are scarce.

**Methods:** By interlinking data from national electronic registries, covering nearly 99% of the Greek population (approximately 11,000,000), between March 2020 and February 2021, when vaccination became available, we recorded confirmed infections and Covid-19-associated hospitalizations and deaths in essentially all adult patients with RA, AS, PsA, SLE, and SSc under treatment (n=74,970, median age of 67.5, 51.2, 58.1, 56.2, 62.2 years, respectively) and in individually matched (1:5) on age, sex, and region of domicile random comparators from the general population.

**Results:** Binary logistic regression analysis after adjusting for age, sex and biologic agents, revealed that RA, PsA, SLE and SSc, but not AS patients, had significantly higher risk of infection (by 43%, 25%, 20% and 49%, respectively), and hospitalization for Covid-19 (by 81%, 56%, 94%, and 111%, respectively), possibly due, at least in part, to increased testing and lower threshold for admission. Patients with RA and SSc had indeed higher Covid-19 associated mortality rates [OR:1.86 (95% CI 1.37 to 2.52) and OR:2.90 (95% CI 0.97 to 8.67), respectively] compared to the general population. Each additional year of age increased the risk of hospitalization for Covid-19 by 3% (OR 1.030, 95% CI: 1.028 to 1.034) and the risk of Covid-19 related death by 8% (OR 1.08, 95% CI: 1.07 to 1.09), independently of gender, systemic rheumatic disease, and biologic agents. A further analysis using AS patients as the reference category, adjusting again for age, sex and use of biologic agents showed that patients with SSc had increased mortality (OR: 6.90, 95% CI: 1.41 to 33.72), followed by SLE (OR: 4.05 95% CI: 0.96 to 17.12) and RA patients (OR: 3.65, 95% CI: 1.06 to 12.54), whereas PsA patients had comparable mortality risk with AS patients.

**Conclusion:** Comparing to the general population, Covid-19 may have a more severe impact in real-world patients with systemic rheumatic disease. Covid-19 related mortality is increased in subgroups of patients with specific rheumatic diseases, especially in older ones, underscoring the need for priority vaccination policies and access to targeted treatments.

## Introduction

Over the last two years, corona virus disease 2019 (Covid-19) has been associated with increased morbidity and mortality in the general population[1]. Evidence, however, regarding systemic rheumatic diseases which affect almost 2% of the population[2] is not conclusive. In the systematic literature review performed to inform the respective European Alliance of Associations for Rheumatology recommendations, no signal for increased mortality in systemic rheumatic diseases compared to the general population was detected[3]. Similar results were presented in a recent meta-analysis examining data from 26 observational studies[4]. On the other hand, a large meta-analysis of 71 studies showed that patients with systemic rheumatic disease displayed increased odds for mortality[5]. The same uncertainty also applies to the question whether systemic rheumatic disease patients are more susceptible to contracting Covid-19, with systematic literature reviews and meta-analyses having discordant results[3–5]. Moreover, the known increased rates of concomitant comorbidities, including arterial hypertension, cardiovascular disease, diabetes, and malignancy[6–12] may have a major additional impact. In any case, whether a propensity of systemic rheumatic disease patients towards more severe Covid-19 associated outcomes, as found in some studies, is true for some or for all diseases still remains unclear.

Herein, we aimed to investigate the Covid-19-associated risk of infection, hospitalization, and death in patients with rheumatoid arthritis (RA), ankylosing spondylitis (AS), psoriatic arthritis (PsA), systemic lupus erythematosus (SLE) and systemic sclerosis (SSc) in comparison with the general population, by interlinking data from national electronic registries between March 2020, when Covid-19 pandemic started in Greece, and February 2021, when vaccination became available to patients with systemic rheumatic diseases.

## Methods

### Setting

The first confirmed case of Covid-19 in Greece was identified on 26 February 2020. Until 28 February 2021 there had been 191,100 confirmed cases and 6,504 Covid-19 associated deaths reported in the country. During the first year of the pandemic several measures were implemented to contain transmission of the virus, including a strict lock-down from 23 March 2020 until 14 May 2020; schools and tourist enterprises did not reopen until 1 June 2020 and 1 July 2020, respectively. A second pandemic wave in November 2020 led to a new lock-down from 7 November 2020, with measures remaining in place until after March 2021 in some regions, including Athens, the country’s capital. On the other hand, priority vaccination against Covid-19 in Greece became available for patients with systemic rheumatic disease in mid-March 2021.

### Data Sources

In Greece, an electronic database for social security services (IDIKA) operates since 2011, currently covering 99% of the country’s population of about 11,000,000 people. Information about prescribed medications, medical diagnoses (based on the specific International Classification of Disease [ICD-10]), age, gender and region of domicile derived from this database can be linked to the nationwide death records as well as to the national Covid-19 digital registry, which includes data on hospitalizations and deaths of all confirmed Covid-19 cases in the country.

### Study Population

In this nationwide, population-based cohort study we identified all adult patients with RA, AS, PsA, SLE, and SSc alive on 1-March-2020 and matched each patient to 5 random referents from the general population for gender, age and region of domicile. For this purpose, we used our published data[13] derived from the electronic prescription database for social security services (IDIKA) including all adult (aged ≥18years old) patients who had filled at least one prescription for corticosteroids, conventional synthetic disease modifying anti-rheumatic drugs (DMARDs), immunosuppressants, biologic DMARDs, targeted synthetic DMARDs, advanced vasodilatory medications or antifibrotic agents between 1-January-2015 and 31-December-2019 with a diagnosis of either RA, AS, PsA, SLE or SSc, based on prespecified for each disease ICD-10 codes. To avoid selection bias, we limited our requirements for inclusion in the cohort to one filled prescription with any of the above-mentioned medications of interest between 2015-2019. Therefore, only a very small minority of patients consistently buying their rheumatology drugs over the counter for 5 consecutive years would not have been captured.

For all subjects in our cohort, we retrieved data on age, gender, use of biologic agents, and by crosslinking with the national Covid-19 registry, data on SARS-Cov-2 infection confirmed by reverse-transcriptase-polymerase-chain-reaction or antigen-detecting rapid diagnostic testing, as well as Covid-19-associated hospitalization and death, during the study period. Following approval of our formal request, the Greek Ministry of Health granted us permission to use anonymized data deposited in the social security services (IDIKA) database and the National Covid-19 digital registry, according to the European legislation for General Data Protection Regulation, (27 April 2016) and the Greek national laws (4600/2019, 4624/2019, 3892/10, 3418/2005).

### Statistical Analysis

Continuous variables were presented as mean (standard deviation [SD]) if having a normal distribution and as median (Q1-Q3) if non-normal. Categorical variables were reported as numbers and percentages. Multivariable binary logistic regression analysis adjusted for age, sex, presence of RA, AS, PsA, SLE and SSc and use of biologic agents was used to estimate Odds Ratios (OR) for a. SARS-CoV-2 infection, b. Covid-19 associated hospitalization and c. Covid-19-related death separately for RA, AS, PsA, SLE and SSc patients and collectively for patients with inflammatory arthritis (RA, AS and PsA) and for all patients with systemic rheumatic disease, each time using the general population as comparison group. A further binary logistic regression analysis to estimate ORs for Covid-19 related hospitalization and death, included only SARS-CoV-2 infected RA, PsA, SLE and SSc patients using SARS-CoV-2 infected AS patients as reference category. These models were adjusted for the same covariates as mentioned above. In all analyses age was treated as a continuous variable, increasing by year. There were no missing data on covariates we selected to record. The level of statistical significance was set at a p-value of ≤0.05. Statistical analysis was performed using the Stata statistical software package (StataCorp. 2013. Stata Statistical Software: Release 13. College Station, TX: StataCorp LP.).

## Results

### Cohort demographics

Our search of the IDIKA database, using the above-described criteria, retrieved 40,014 RA patients (79% female), 9,566 AS patients (43% female),13,405 PsA patients (55% female), 9,960 SLE patients (90% female) and 2,025 SSc patients (88% female). RA patients were older compared to other systemic rheumatic disease patients with a median(Q1-Q3) age of 67.5 (57.4-76.0) years, followed by SSc [62.2 (51.7-71.0) years], PsA [58.1 (48.6-68.0) years], SLE [56.2 (45.2-67.2) years] and AS patients [51.2 (41.9-60.4) years]. Total follow-up was 74,765 person-years for systemic rheumatic disease patients and 372,019 person-years for their matched comparators. **Table 1** shows demographics of systemic rheumatic disease patients and their matched referents from the general population. SARS-CoV-2 infections, Covid-19 related hospitalizations and deaths occurring in our cohort between 1 March 2020 and 28 February 2021 are also shown in **Table 1**.

**Table 1.**
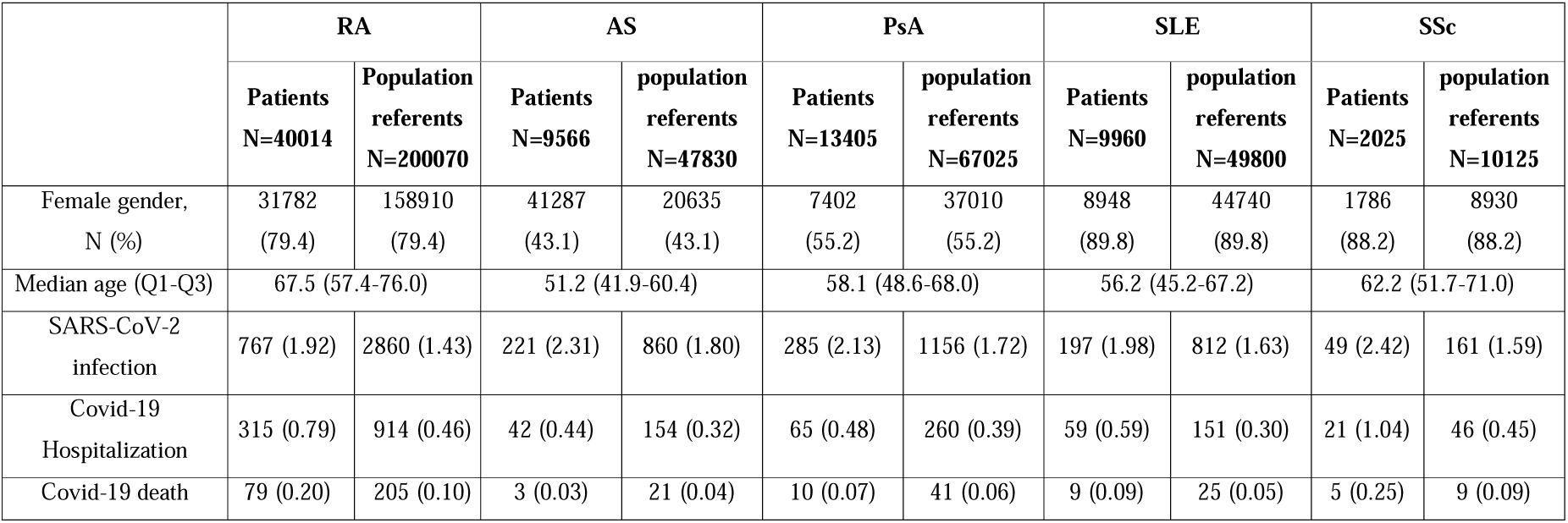
Demographics and number of SARS-CoV-2 infections, number of Covid-19-associated hospitalizations and deaths, and number of deaths from all causes recorded between 1-3-2020 and 28-2-2021 among patients with rheumatoid arthritis (RA), ankylosing spondylitis (AS), psoriatic arthritis (PsA), systemic lupus erythematosus (SLE), systemic sclerosis (SSc) and their matched population referents.

|  | **RA** |  | **AS** |  | **PsA** |  | **SLE** |  | **SSc** |  |
| --- | --- | --- | --- | --- | --- | --- | --- | --- | --- | --- |
|  | **Patients**<br>**N=40014** | **Population**<br>**referents**<br>**N=200070** | **Patients**<br>**N=9566** | **population**<br>**referents**<br>**N=47830** | **Patients**<br>**N=13405** | **population**<br>**referents**<br>**N=67025** | **Patients**<br>**N=9960** | **population**<br>**referents**<br>**N=49800** | **Patients**<br>**N=2025** | **population**<br>**referents**<br>**N=10125** |
| Female gender,<br>N (%) | 31782<br>(79.4) | 158910<br>(79.4) | 41287<br>(43.1) | 20635<br>(43.1) | 7402<br>(55.2) | 37010<br>(55.2) | 8948<br>(89.8) | 44740<br>(89.8) | 1786<br>(88.2) | 8930<br>(88.2) |
| Median age (Q1-Q3) | 67.5 (57.4-76.0) |  | 51.2 (41.9-60.4) |  | 58.1 (48.6-68.0) |  | 56.2 (45.2-67.2) |  | 62.2 (51.7-71.0) |  |
| SARS-CoV-2<br>infection | 767 (1.92) | 2860 (1.43) | 221 (2.31) | 860 (1.80) | 285 (2.13) | 1156 (1.72) | 197 (1.98) | 812 (1.63) | 49 (2.42) | 161 (1.59) |
| Covid-19<br>Hospitalization | 315 (0.79) | 914 (0.46) | 42 (0.44) | 154 (0.32) | 65 (0.48) | 260 (0.39) | 59 (0.59) | 151 (0.30) | 21 (1.04) | 46 (0.45) |
| Covid-19 death | 79 (0.20) | 205 (0.10) | 3 (0.03) | 21 (0.04) | 10 (0.07) | 41 (0.06) | 9 (0.09) | 25 (0.05) | 5 (0.25) | 9 (0.09) |

### Comparison of patients with RA, AS, PsA, SLE and SSc with matched population referents

**Table 2** shows incidence rates per 1000 patient-years for Covid-19 associated infection, hospitalization, and death among patients with RA, AS, PsA, SLE and SSc, and their matched population comparators. Rates of infection were higher among patients with any chronic inflammatory arthritis, rates of hospitalization were higher among patients with RA, SLE and SSc, whereas only RA patients had higher mortality rates comparing to the general population.

**Table 2.** Incidence Rates (IR) per 1000 person years with 95% Confidence Intervals (CI) for Covid-19 infection, hospitalization and death among rheumatoid arthritis (RA), ankylosing spondylitis (AS), psoriatic arthritis (PsA), systemic lupus erythematosus (SLE), and systemic sclerosis (SSc) patients and their matched referents from the general population.

|  | **RA** |  | **AS** |  | **PsA** |  | **SLE** |  | **SSc** |  |
| --- | --- | --- | --- | --- | --- | --- | --- | --- | --- | --- |
|  | patients | controls | patients | controls | patients | controls | patients | controls | patients | controls |
| Covid-19 infection<br>IR (95% CI) | **19.2**<br>**(17.9-20.7)** | **14.4**<br>**(13.9-15.0)** | **23.1**<br>**(20.3-26.4)** | **18.0**<br>**(16.9-19.3)** | **21.3**<br>**(19.0-23.9)** | **17.3**<br>**(16.4-18.4)** | 19.8<br>(17.2-22.8) | 16.4<br>(15.3-17.6) | 24.3<br>(18.4-32.2) | 16.0<br>(13.7-18.7) |
| Covid-19 Hospitalization<br>IR (95% CI) | **7.9**<br>**(7.1-8.8)** | **4.6**<br>**(4.3-4.9)** | 4.4<br>(3.2-5.9) | 3.2<br>(2.8-3.8) | 4.9<br>(3.8-6.2) | 3.9<br>(3.5-4.4) | **5.9**<br>**(4.6-7.7)** | **3.0**<br>**(2.6-3.6)** | **10.4**<br>**(6.8-16.0)** | **4.6**<br>**(3.4-6.1)** |
| Covid-19 Deaths<br>IR (95% CI) | **1.98**<br>**(1.59-2.47)** | **1.04**<br>**(0.90-1.19)** | 0.31<br>(0.10-0.97) | 0.44<br>(0.29-0.68) | 0.75<br>(0.40-1.39) | 0.62<br>(0.45-0.84) | 0.91<br>(0.47-1.74) | 0.50<br>(0.34-0.75) | 2.48<br>(1.03-5.96) | 0.89<br>(0.47-1.72) |

After adjusting for age, sex and the use of biologic agents, binary logistic regression analysis (**Table 3**) revealed that patients with RA had higher probability to contract SARS-Cov-2 by 43% (OR 1.43, 95% CI: 1. 29 to 1.57), to get hospitalized by 81% (OR 1.81, 95% CI: 1.55 to 2.10) and to die of covid-19 by 86% (OR 1.86, 95% CI: 1.37 to 2.52) vs the general population. As far as AS patients were concerned, these displayed comparable to the general population risk of infection, hospitalization, or death due to Covid-19. A higher risk of Covid-19 infection and hospitalization, but not death was found for PsA patients (OR 1.25, 95% CI: 1.02 to 1.53 for infection and OR 1.56 95% CI: 1.08 to 2.24 for hospitalization, respectively), compared to their matched referents from the general population. Collectively, patients with chronic inflammatory arthritis (n=62,985) had increased risk of infection by 39%, (OR 1.39, 95% CI: 1.27 to 1.51), of hospitalization by 74% (OR 1.74, 95% CI: 1.52 to 2.00) and of death due to Covid-19 by 74% (OR 1.74, 95% CI: 1.31 to 2.32).

**Table 3.** Odds Ratio with 95% Confidence Interval for Covid-19 associated infection, hospitalization, and death among all patients with Rheumatoid Arthritis (RA), Ankylosing Spondylitis (AS), Psoriatic Arthritis (PsA), Systemic Lupus Erythematosus (SLE), Systemic Sclerosis (SSc) and their matched referents from the general population. Binary logistic Regression analysis adjusted for age, gender and use of biologics.

| **A. Binary logistic regression analysis including all RA, AS, PsA, SLE, SSc patients and matched population referents** |  |  |  |  |  |
| --- | --- | --- | --- | --- | --- |
|  | **RA** | **AS** | **PsA** | **SLE** | **SSc** |
| **Covid-19 Infection**<br>**OR (95%CI)** | **1.43 (1.29 to 1.57)** | 1.31 (0.90 to 1.90) | **1.25 (1.02 to 1.53)** | **1.20 (1.02 to 1.40)** | **1.49 (1.07 to 2.08)** |
| **Covid-19**<br>**Hospitalization OR**<br>**(95%CI)** | **1.81 (1.55 to 2.10)** | 1.02 (0.40 to 2.60) | **1.56 (1.08 to 2.24)** | **1.94 (1.43 to 2.64)** | **2.11 (1.24 to 3.62)** |
| **Covid-19 Death**<br>**OR (95%CI)** | **1.86 (1.37 to 2.52)** | 0.33 (0.1 to 8.54) | 1.23 (0.52 to 3.23) | 1.60 (0.73 to 3.50) | 2.90 (0.97 to 8.67) |

Regarding patients with SLE, higher risks of Covid-19 infection and hospitalization (OR 1.20, 95% CI: 1.02 to 1.40 for infection, and OR 1.94, 95% CI: 1.43 to 2.63 for hospitalization, respectively), but not of death, were noted. Similarly, patients with SSc had higher risks of infection and hospitalization comparing to the general population (OR 1.49, 95% CI: 1.07 to 2.08 for infection and OR 2.11, 95% CI: 1.24 to 3.62 for hospitalization, respectively). The risk of Covid-19 associated death was also higher in SSc patients (OR 2.90, 95% CI: 0.97 to 8.67, p=0.057) compared to the general population, albeit with marginal significance due to lower numbers.

### Comparison of rheumatic disease patients collectively with matched population referents

The total of 74,970 patients [median (Q1-Q3) age 62.2 (51.2-72.6) years, 72% females) was compared to the total of individually matched (1:5) on age, sex, and region of domicile random referents from the general population. SARS-Cov-2-related infection rates, hospitalization rates and death rates collectively for patients with systemic rheumatic disease were 20.32 (95% CI 19.32 to 21.36), 6.71 (95% CI 6.15 to 7.33) and 1.42 (95% CI 1.17 to 1.72) per 1000 person-years, respectively. The relevant figures for their matched referents were 15.72 (95% CI 15.32 to 16.13), 4.10 (95% CI 3.90 to 4.31) and 0.81 (95% CI 0.72 to 0.91) per 1000 person-years, respectively.

As depicted in **Figure 1**, binary logistic regression analysis performed in the entire cohort revealed that patients collectively, had 34% higher risk to get infected (OR 1.34, 95% CI: 1.24 to 1.44), 79% higher risk to get hospitalized (OR 1.79, 95% CI: 1.59 to 2.02) and 77% higher risk to die (OR 1.77, 95% CI: 1.36 to 2.30), compared to their matched referents from the general population.

**Figure 1.**
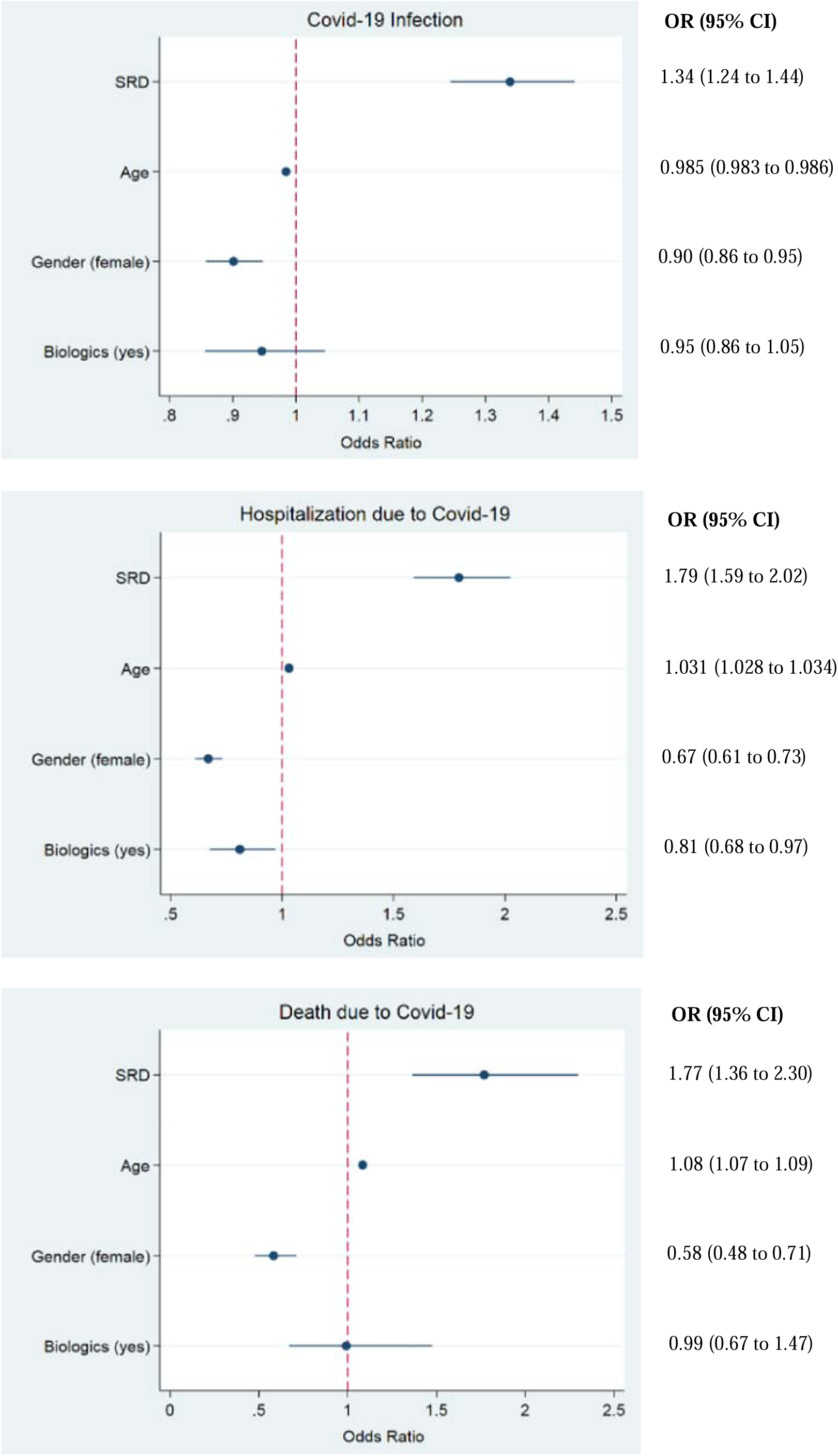
Odds Ratio with 95% Confidence Intervals for SARS-CoV-2 infection (upper panel), hospitalization due to Covid-19 (middle panel) and death due to Covid-19 (lower panel) between 1-3-2020 and 28-2-2021 in the entire cohort of patients with systemic rheumatic disease (SRD) and their matched comparator subjects from the general population. Binary logistic regression model adjusted for presence of SRD, age, gender and use of biologic agents.

Each additional year of age increased the risk of hospitalization for Covid-19 by 3% (OR 1.030, 95% CI: 1.028 to 1.034) and the risk of Covid-19 related death by 8% (OR 1.08, 95% CI: 1.07 to 1.09), independently of gender, systemic rheumatic disease, and biologic agents.

Female gender had a protective effect against infection, hospitalization, and death from SARS-CoV-2 by 10% (OR 0.90, 95% CI: 0.86 to 0.95), 33% (OR 0.67, 95% CI: 0.61 to 0.73) and 42% (OR 0.58, 95% CI: 0.48 to 0.71) respectively, regardless of age, presence of systemic rheumatic disease and use of biologics.

Finally, after adjusting for age, sex, and the presence or not of systemic rheumatic disease, use of biologic agents did not significantly affect infection rates or Covid-19 associated mortality but exerted a protective effect against hospitalization for Covid-19 by 19% (OR 0.81, 95% CI: 0.68 to 0.97) (**Figure 1**).

### Subgroup analyses in patients with Covid-19

To investigate for potential differences between systemic rheumatic diseases we performed a subgroup analysis within patients infected with SARS-Cov-2, using AS patients as the reference category. After adjustment for age, sex and biologic agents we found that patients with SSc had worse prognosis regarding both Covid-19 associated hospitalization and death (OR: 2.60, 95% CI: 1.21 to 5.56 for hospitalization, OR: 6.90, 95% CI: 1.41 to 33.72 for death), followed by SLE (OR: 1.96, 95% CI: 1.12 to 3.43 for hospitalization, OR: 4.05 95% CI: 0.96 to 17.12 for death), and by RA patients (OR: 1.53, 95% CI: 0.99 to 2.37 for hospitalization, OR: 3.65, 95% CI: 1.06 to 12.54 for death), whereas PsA patients did not have significantly higher risk of hospitalization and death in comparison with AS patients **(Figure 2)**.

**Figure 2.**
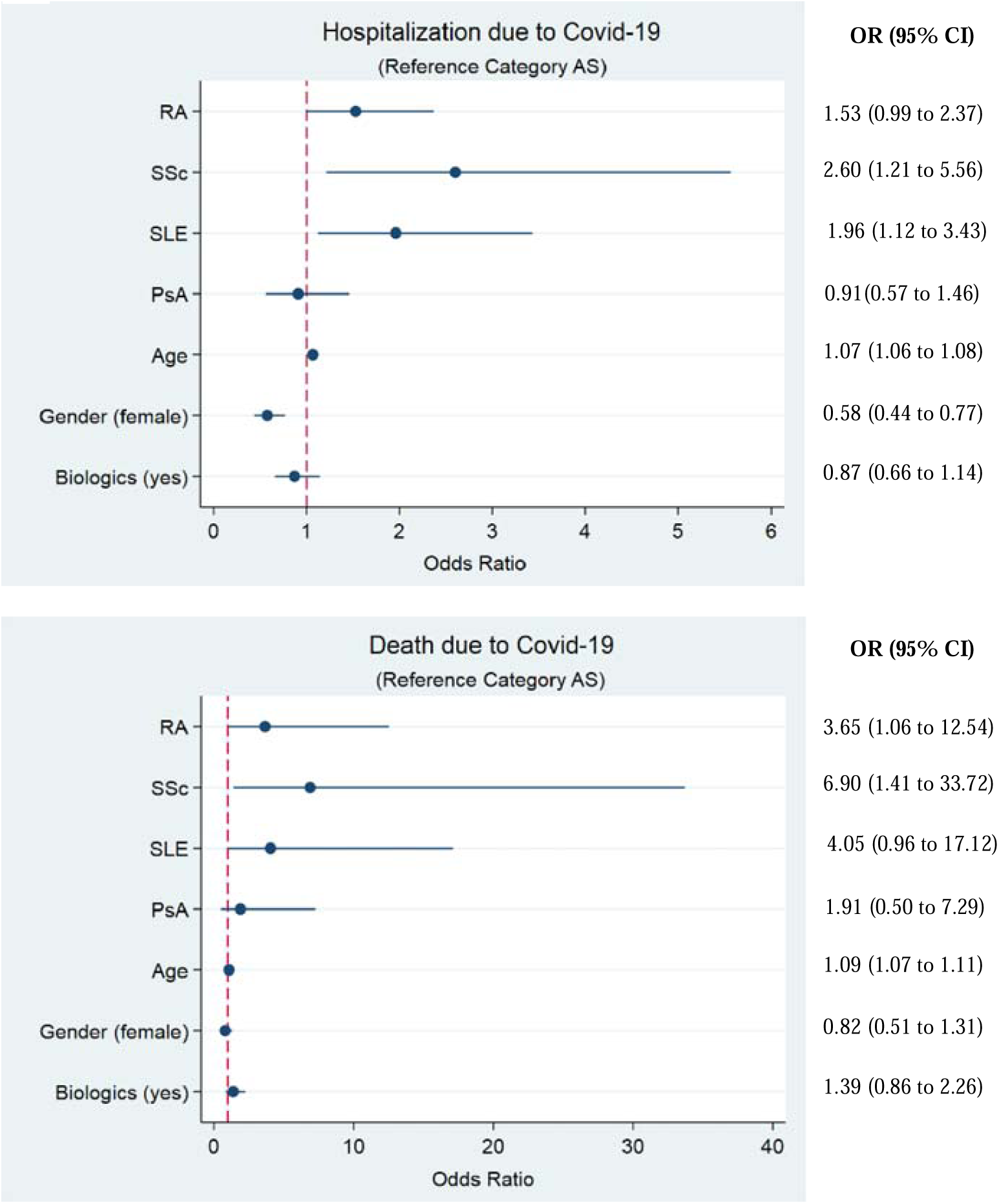
Odds Ratio with 95% Confidence Intervals for hospitalization and death due to Covid-19 in the subgroup of patients with systemic rheumatic disease (SRD) infected with SARS-CoV-2, between 1-3-2020 and 28-2-2021. Model adjusted for underlying SRD, age, gender and use of biologic agents. Reference category: Ankylosing spondylitis (AS). RA: Rheumatoid Arthritis, SSc: Systemic Sclerosis, SLE: Systemic Lupus Erythematosus and PsA: Psoriatic Arthritis.

## Discussion

Our 12-month study, analysing data derived from the inter-linkage of large nationwide databases covering the entire Greek population, shows that unvaccinated patients with RA, PsA, SLE and SSc, but not AS, have a higher risk of SARS-Cov-2 infection and hospitalization due to Covid-19 compared to the general population. Patients with RA and SSc had also higher Covid-19 associated mortality rates compared to the general population. The analysis of the entire cohort revealed an inverse correlation between age and the risk of SARS-Cov-2 infection, which is not surprising because of the increased protection measures taken by older subjects. Notably, in this same analysis each additional year of age increased the risk of Covid-19-related death by 8%, irrespective of gender and disease. Therefore, since we were not able to correct for comorbidities which are known to be increased in any systemic rheumatic disease, the higher mortality in RA and SSc patients, who are older than patients with AS, PsA and SLE, could be partly explained by the fact that the burden of comorbidities could have a synergistic effect on Covid-19 outcomes with advanced age.

To the best of our knowledge only one previous nation-wide study assessing mortality risk of patients with chronic inflammatory arthritis versus the general population has been published[14]. This was a 6-month study in Sweden, a unique country where public measures were far less strict, especially during the first pandemic wave, compared to the rest of the world. The authors reported that death rates were increased when adjusting only for age, sex and region of domicile, but this risk was mitigated when additionally adjusting for comorbidities and socioeconomic factors. The adjusted HR for Covid-19 associated death in all inflammatory joint diseases examined in this study was 1.18 (95% CI 0.97 to 1.44), while the adjusted HR for Covid-19 associated death in RA was 1.27 (1.02 to 1.59), compared with matched general population comparator subjects, which is significantly higher, indeed. The authors concluded that, in absolute terms, risks of serious outcomes from Covid-19 in patients with inflammatory joint disease are strongly affected by age and the presence of comorbidities. Our findings, in accordance with the Swedish nation-wide study, also indicate an increased likelihood of death due to Covid-19 among RA patients in comparison to the general population, but no increased risk among the other disease groups examined, with the exception of SSc patients, for which Covid-19 associated mortality was also higher compared to the general population.

A second nation-wide 5-month study from South Korea included 133,609 adult subjects tested for SARS-COV-2 (3.65% tested positive), of whom 8,297 had inflammatory rheumatic diseases. In comparison to the general population, patients with rheumatic disease had a 19% increased likelihood of testing positive for SARS-COV-2, 26% higher risk of severe Covid-19 outcomes and 69% higher risk of Covid-19 associated death. Patients with rheumatic diseases receiving higher corticosteroid doses (≥10 mg prednisolone per day) were particularly vulnerable to developing worse Covid-19 associated outcomes[15]. Notably, our results are in line with this study, as well as with the results of a meta-analysis by Conway et al. reporting an OR of 1.74 (95%CI: 1.08 to 2.80) for Covid-19-related death in systemic rheumatic disease patients in comparison with the general population[5]. It should be here noted that both our study and the meta-analysis of Conway et al. could not adjust for comorbidities, disease severity or disease duration, due to the lack of the corresponding data in the data sources used. This important limitation should be taken in account when interpreting these findings, given the high frequency of comorbidities among patients with systemic rheumatic disease[6–12] and the strong influence that comorbidities and disease activity exert on Covid-19 associated outcomes in this patient population[16, 17].

Regarding the risk of Covid-19 infection, with the exception of AS patients, all other patient subgroups were at higher risk, in agreement also with a general population-based cohort study[18]and two recent-metanalyses[4, 5]. However, the possibility that these patients were more susceptible to increased rates of testing cannot be excluded. Therefore, whether a tendency to contract the infection more easily, together with the increased comorbidity burden, can explain the higher Covid-19 associated death risk found collectively in systemic rheumatic disease patients compared to the general population needs further study[19].

As regards to hospitalization rates, there are three nation-wide studies from Sweden[14], Denmark[20] and Iceland[21], in line with our findings, while two meta-analyses[4, 5] report different results on this matter. This could be interpreted in light of several differences between countries, such as local guidelines, intensity of pandemic wave, saturation level of health care system, access to healthcare facilities and other confounders[22]. Increased hospitalization rates could be possibly also due, at least in part, to lower threshold for admission of patients suffering from a systemic rheumatic disease. Besides, in concert with our results various factors have been previously identified to associate with Covid-19-related hospitalization, including male gender, higher age and specific diseases like RA, vasculitis, and connective tissue diseases[17, 20, 23–26]. Importantly, in accordance with our observations, treatment with biologic agents has been negatively associated with hospital admission in other studies, as well [4, 24, 27].

In general, findings about Covid-19 related outcomes in systemic rheumatic disease patients should be interpreted with caution. Inconsistencies between various studies may pertain to ethnic or racial differences[28], to the heterogeneity of systemic rheumatic disease and their treatment, differences in the intensity of pandemic waves and the capacity of health care system, but also to the different study designs or timing of data acquisition with regard to the implementation of patient vaccination policies. That said, we should note that our study was performed before vaccination was available among systemic rheumatic disease patients in Greece.

A recently published study also raised some concerns about high rate of biases (e.g participation and ascertainment bias) occurring in the studies being published about this topic[29]. This was also noted in a recent systematic literature review performed to inform respective EULAR recommendations[3]. In the strengths of our study, one can include the following: firstly, it is a nationwide study examining the whole population of our country (approximately 11 million people) during the whole first year of the pandemic. To further strengthen our methodology, matching with the general population was adjusted for area of residence, limiting biases concerning access to healthcare facilities and regional differences in Covid-19 incidence rate. Thus, selection bias occurring in registries is likely eliminated. Also, in contrast to other studies, we have examined simultaneously five major systemic rheumatic disease in the same population which also allowed us to make comparison between diseases. Indeed, our data show that patients with SSc have the worse Covid-19-outcomes, followed by SLE and RA, PsA and AS.

One major weakness of our study, as mentioned above, is the inability to adjust our findings for disease duration, disease activity and the presence of comorbidities. However, in the sense that comorbidities now tend to be seen as an inherent component of systemic rheumatic disease, we believe that it is hard to decipher whether the higher Covid-19 associated death rate observed in certain of these patient’ subgroups should be attributed to the systemic rheumatic disease per se or to the concomitant presence of other diseases.

## Conclusions

The risk of death from Covid-19 was found higher collectively for all systemic rheumatic disease patients versus matched referents from the general population, implying that priority vaccination policies and access to targeted therapeutic approaches are important, especially for older patients. The increased death rate is probably attributable to the presence of comorbidities, which are inherent to the chronic inflammatory process characterizing systemic rheumatic diseases. However, Covid-19 outcomes (including death) clearly differ between patients with RA, AS, PsA, SLE and SSc. Further studies could help in effective risk stratification for systemic rheumatic disease patients and therefore result in better outcomes.

## Supporting information

STROBE checklist

## Data Availability

The data that support the findings of this study are available from the Data Protection Officer of the Greek Ministry of Health but restrictions apply to the availability of these data, which were used under license for the current study, and so are not publicly available. Data are however available from the authors upon reasonable request and with permission of the Data Protection Officer of the Greek Ministry of Health.

## List of abbreviations

AS: Ankylosing Spondylitis
CI: Confidence Interval
Covid-19: Corona virus disease 2019
DMARDs: Disease Modifying Anti-Rheumatic Drugs
EULAR: European League Against Rheumatism
ICD-10: International Classification of Disease-10
OR: Odds Ratio
PsA: Psoriatic Arthritis
RA: Rheumatoid Arthritis
SARS-CoV-2: Severe Acute Respiratory Syndrome Coronavirus 2
SD: Standard Deviation
SLE: Systemic Lupus Erythematosus
SRD: Systemic Rheumatic Disease
SSc: Systemic Sclerosis

## Declarations

### Ethics approval and consent to participate

The Data Protection Office of the Greek Ministry of Health (17 Aristotelous str, 10187, Athens, Greece), gave ethical approval for this work, granting permission for the use of anonymized data deposited in the social security services (IDIKA) database and the national Covid-19 digital registry, according to the European legislation for General Data Protection Regulation, (27 April 2016) and the Greek national laws (4600/2019, 4624/2019, 3892/10, 3418/2005).

### Patient and Public Involvement statement

Patients or the public WERE NOT involved in the design, or conduct, or reporting, or dissemination plans of our research.

### Consent for publication

Not applicable

### Competing interests

None Declared

### Funding

This study was partly supported by the Kleon Tsetis Foundation, in the context of promoting academic research initiatives within its COVID-19 response framework

### Authors’ contributions

VKB contributed to study design, curation of data and drafting the manuscript, GEF contributed to study design and drafting the manuscript, IT, KM, AT and PM contributed to data curation, GK contributed to statistical analysis, GV and DP contributed to study design and statistical analysis, MT contributed to study design and critically revised the manuscript, PPS conceived the original idea, supervised the project and contributed to study design, writing and reviewing the manuscript.

## Acknowledgements

Not applicable

## Notes

### Competing Interest Statement

The authors have declared no competing interest.

### Author Declarations

The Data Protection Office of the Greek Ministry of Health (17 Aristotelous str, 10187, Athens, Greece, /), gave ethical approval for this work, granting permission for the use of anonymized data deposited in the social security services (IDIKA) database and the national Covid-19 digital registry, according to the European legislation for General Data Protection Regulation, (27 April 2016) and the Greek national laws (4600/2019, 4624/2019, 3892/10, 3418/2005).

